# IL-2-mediated CD4 T-cell activation correlates highly with effective serological and T-cell responses to SARS-CoV-2 vaccination in people living with HIV (PLWH)

**DOI:** 10.1101/2024.05.28.24308045

**Authors:** Akshita Gupta, Elda Righi, Angelina Konnova, Concetta Sciammarella, Gianluca Spiteri, Vincent Van Averbeke, Matilda Berkell, An Hotterbeekx, Assunta Sartor, Massimo Mirandola, Surbhi Malhotra-Kumar, Anna Maria Azzini, Diletta Pezzani, Maria Grazia Lourdes Monaco, Guido Vanham, Stefano Porru, Evelina Tacconelli, Samir Kumar-Singh

## Abstract

People living with HIV (PLWH) despite having appreciable depletion of CD4^+^ T-cell show a good SARS-CoV-2 vaccination response. The underlying mechanism(s) are currently not understood. We studied serological and polyfunctional T-cell responses in PLWH receiving anti-retroviral therapy stratified on CD4^+^ counts as PLWH-high (CD4 ≥500 cells/μL) and PLWH-low (<500 cells/μL). Responses were assessed longitudinally before the first vaccination (T0), 1-month after the first dose (T1), and 3- months (T2), and 6-months (T3) after the second dose. Expectedly, both PLWH-high and -low groups developed similar serological responses after T2, which were also non-significantly different to age and vaccination-matched HIV-negative controls at T3. The IgG titers were also protective showing a good correlation with ACE2-neutralizations (R=0.628, P=0.005). While no difference at T3 was observed between PLWH and controls in activated CD4^+^CD154^+^ and CD4^+^ memory T-cells, spike-specific CD4^+^ polyfunctional cytokine expression analysis showed that PLWH preferentially express IL-2 (P<0.001) and controls, IFN-γ (P=0.017). CD4^+^ T-cell counts negatively correlated with IL-2- expressing CD4^+^ T-cells including CD4^+^ memory T-cells (Spearman ρ: -0.85 and -0.80, respectively; P<0.001). Our results suggest that the durable serological and CD4^+^ T-cell responses developing in vaccinated PLWH are associated with IL-2-mediated CD4^+^ T-cell activation that likely compensates for CD4^+^ T-cell depletion in PLWH.

## Introduction

The protection provided by mRNA vaccinations is unequivocal among healthy recipients, showing effectiveness for hospitalization up to 92% and 91% for mortality related to COVID-19 infection ^1–3^. Nevertheless, vaccine effectiveness, which is usually assessed using serological methods, is known to wane over time and to be lower against newer SARS-CoV-2 variants including Omicron ^1^. In populations with impaired immunity, including people living with HIV (PLWH), the threat of COVID- 19 adds an additional layer of vulnerability ^4^, as a subset of PLWH despite receiving anti-retroviral therapy, have appreciably lowered number of CD4^+^ T-cells. As CD4^+^ T-cells are necessary for both cellular and humoral immunity, PLWH with lower CD4 counts are expected to have dysregulated immunity toward SARS-CoV-2, thereby hampering antiviral responses and diminishing T-cell cross-recognition and development of immunological memory ^5^.

However, despite these potential threats, most studies have demonstrated good seroconversion rates in PLWH although select subjects displaying CD4^+^ T-cell counts <200 cells/mm^3^ tend to show poorer seroconversion rates ^6–12^. Moreover, studies on cellular immune responses following SARS-CoV-2 vaccination in PLWH have not found any appreciable differences between healthy controls and PLWH ^13,14^. The underlying reasons for this robust humoral and cellular immunity in PLWH remain unknown. Additional concerns in PLWH are also on durability of T-cell immunity, especially memory CD4^+^ T cells, induced by SARS-CoV-2 vaccines. Only a limited number of studies have provided longitudinal T-cell immunophenotyping data related to SARS-CoV-2 vaccination in PLWH^15,16^ and the role of any altered quality of T-cell responses has not been fully explored.

At the current stage of post-COVID-19 pandemic period, heterologous combinations of vaccination and exposure to viral antigens are also expected to shape a distinctive immunological memory ^17,18^. With the development of bivalent (BA.1 or BA.4/5 and XBB1.5) mRNA vaccines, there is hope that an appreciably better response could be seen compared to those with only the ancestral SARS-CoV-2 (Wuhan-Hu-1) monovalent mRNA vaccination. However, antigenic imprinting has been identified as a substantial limitation to the development of an effective secondary adaptive immune response^19^. Thus, achieving and maintaining an optimal vaccine-induced T-cell response remains of particular importance for PLWH.

This study investigates longitudinal dynamics of SARS-CoV-2-specific T-cells before and after primary vaccination, specifically focusing on any altered quality of T-cell responses that may shape humoral and cellular immunity and T-cell memory responses in PLWH population.

## Results

### Demographic and clinical characteristics

Fifty-six participants, 30 PLWH and 26 HIV-negative control individuals working as health care workers, were enrolled in the study during April–December 2021 (**SI-Figure 1**). The median age of PLWH was 54 years (interquartile range (IQR): 45-60) and 55 years for controls (IQR: 47-60) years (IQR: 47-60) years(see **Table 1** for demographic information). All participants received the BNT162b2 vaccine with a schedule of two doses of 30 μg 21±7 days apart. Five controls were reported to have previously been infected with PCR-confirmed SARS-CoV-2. Among PLWH, no previous history of SARS-CoV-2 infection was reported, however, one PLWH was likely infected based on the high anti-Nucleocapsid (N) and anti-Spike (S) titers at baseline (see next section). PLWH in our cohort were on ART and had HIV-RNA levels of <200 copies/mL and therefore considered virally-suppressed; except for one PLWH who consistently had HIV-RNA levels of >400[l]000 copies/mL (patient H173). The median absolute CD4^+^ T-cell count for PLWH at T0 timepoint was 706 cells/mm^3^ (IQR: 349-1007) and remained stable over time, with 3 patients displaying CD4^+^ counts of <200 cells/mm^3^, including patient H173 who had average CD4^+^ counts of 9 cells/mm^3^ during the study. Based on CD4^+^ T-cell counts at baseline (before vaccination), PLWH were stratified as PLWH-high (≥500/mm^3^; n=20) and PLWH-low (<500 cells/mm^3^; n=10). While vaccinations are shown to activate HIV transcription and virion production both in HIV patients as well as in elite controllers who are able to control viral replication without ART^20,21^, no overall alteration in viral load was detected in our PLWH cohort over the studied timepoints. These data suggest that SARS-CoV-2 vaccination does not adversely affect HIV disease status in ART-treated PLWH.

**Figure 1.**
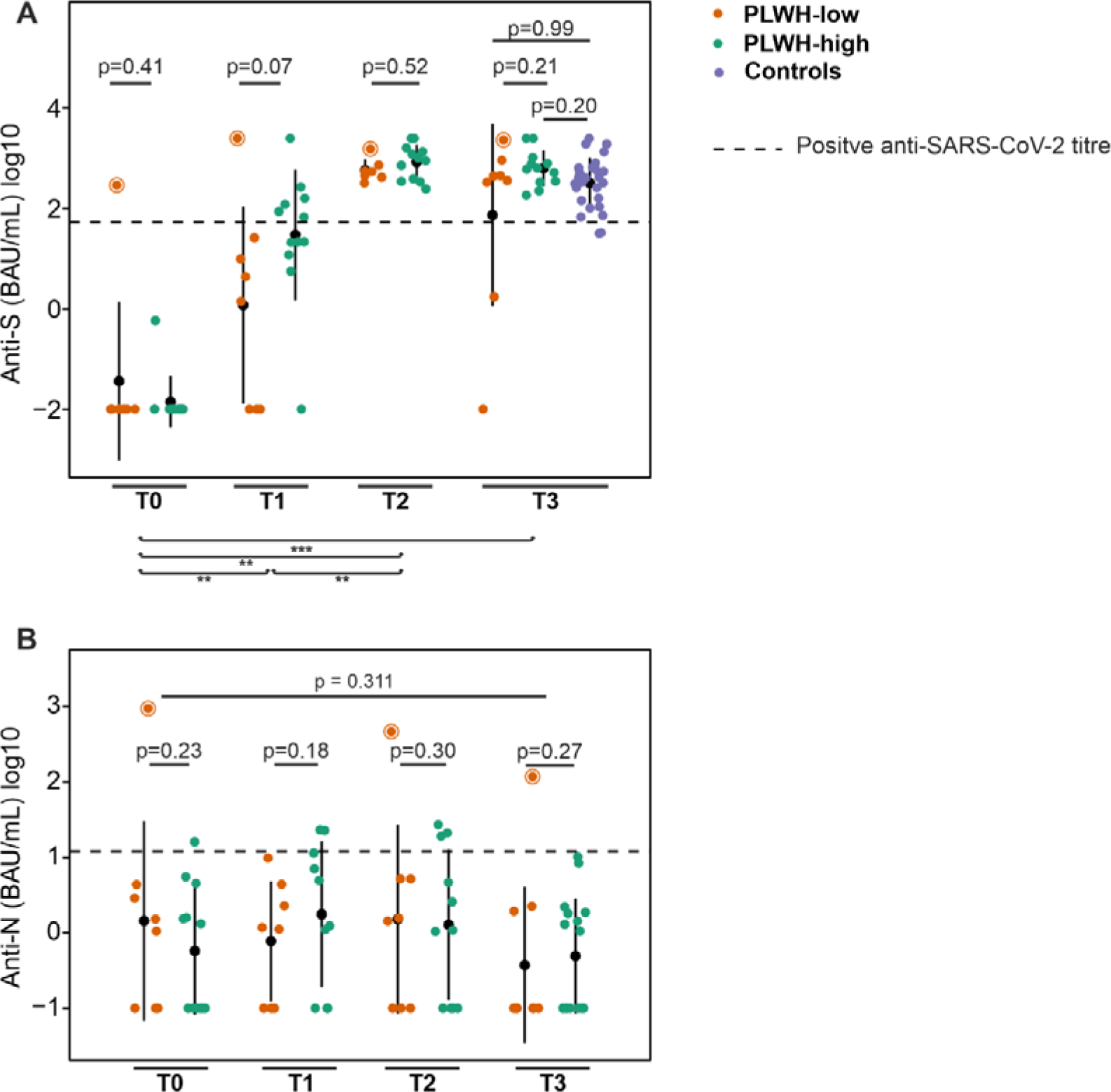
Anti-SARS-CoV-2 serological responses in vaccinated PLWH population stratified as PLWH-high and PLWH-low. (A) Anti-RBD (Wuhan Hu-1) IgG titers measured using the Roche Elecsys anti-SARS-CoV-2 immunoassay in paired serum samples of PLWH-high and PLWH-low following vaccine administration. Roche Elecsys detection limit = 2500 BAU/mL. (B) Anti- Nucleocapsid IgG titers measured using MSD assay in paired samples of PLWH-high and PLWH-low. Control samples were not analysed for anti-N IgG. Dot plots indicate mean (black dot) and 1 standard deviation (whiskers) for (A) and (B). The dashed line indicates the positive SARS-CoV-2 titers for each assay. All data points, including outliers, are displayed. **: p < 0.01. ***: p < 0.001. Highest titers for anti-N and anti-S antibodies was noted for one PLWH-low participant who was likely infected before receiving the first dose (circled dot). T0: Pre-vaccination timepoint; T1: 1 month after the first vaccination; T2: 3 months after the second vaccination; T3: 6 months after the second vaccination.

**Table 1.** Demographic details and comorbidities.

| Patient Characteristics | PLWH<br>N = 30 | Controls<br>N = 26 | P-value |
| --- | --- | --- | --- |
| Male, n (%) | 21 (70%) | 10 (38%) | **0.014<sup>‡</sup>** |
| Age (median, IQR) | 54 (45-60) | 55 (47-60) | 0.800<sup>‡</sup> |
| Positive SARS-CoV-2 PCR, n (%) | 0 (%) | 5 (19%) | **0.016<sup>‡</sup>** |
| CD4+ Nadir (median, IQR) | 198 (91-289) | NA | NA |
| HIV RNA (median, IQR) | 0 (0-20) | NA | NA |
| PLWH-high | 20 | NA | NA |
| PLWH-low | 10 | NA | NA |
| **Antiretroviral therapy** |  |  |  |
| Lamivudine, n (%) | 18 (60%) | NA | NA |
| Efavirenz, n (%) | 2 (6.7%) | NA | NA |
| Raltegravir, n (%) | 2 (6.7%) | NA | NA |
| Atazanavir, n (%) | 3 (10%) | NA | NA |
| Cobicistat, n (%) | 4 (13%) | NA | NA |
| Emtricitabine, n (%) | 7 (23%) | NA | NA |
| Tenofovir, n (%) | 8 (27%) | NA | NA |
| Dolutegravir, n (%) | 12 (40%) | NA | NA |
| Rilpivirine, n (%) | 6 (20%) | NA | NA |
| Darunavir, n (%) | 5 (17%) | NA | NA |
| Ritonavir, n (%) | 4 (13%) | NA | NA |
| Nevirapine, n (%) | 1 (3.3%) | NA | NA |
| Doravirine, n (%) | 1 (3.3%) | NA | NA |
| Disoproxil, n (%) | 2 (6.7%) | NA | NA |
| Fumarate, n (%) | 1 (3.3%) | NA | NA |
| Abacavir, n (%) | 4 (13%) | NA | NA |
| **Comorbidities** |  |  |  |
| Hypertension, n (%) | 7 (23%) | 2 (7.7%) | 0.200<sup>#</sup> |
| Immunological diseases requiring immunosuppressive agents, n (%) | 2 (6.7%) | 1 (3.8%) | 0.900<sup>#</sup> |
| Neurological disease, n (%) | 1 (3.3%) | 2 (7.7%) | 0.600<sup>#</sup> |
| Solid organ or hematological cancer, n (%) | 2 (6.7%) | 3 (12%) | 0.700<sup>#</sup> |
| Chronic pulmonary disease, n (%) | 2 (6.7%) | 0 (0%) | 0.500<sup>#</sup> |
| Liver disease, n (%) | 5 (17%) | 1 (3.8%) | 0.200<sup>#</sup> |
| Thyroid disease, n (%) | 1 (3.3%) | 3 (12%) | 0.300<sup>#</sup> |
| Vascular disease, n (%) | 3 (10%) | 0 (0%) | 0.200<sup>#</sup> |
| Ulcer, n (%) | 1 (3.3%) | 0 (0%) | 0.900<sup>#</sup> |
| Diabetes (with or without damage), n (%) | 3 (10%) | 0 (0%) | 0.200<sup>#</sup> |
| Chronic renal failure (with or without need of dialysis), n (%) | 7 (23%) | 1 (3.8%) | 0.059<sup>#</sup> |
| Cardiovascular disease (ischemic/arrythmia), n (%) | 5 (17%) | 3 (12%) | 0.700<sup>#</sup> |
| Gastrointestinal disorder, n (%) | 3 (10%) | 0 (0%) | 0.200<sup>#</sup> |
| Blood disorder, n (%) | 3 (10%) | 0 (0%) | 0.200<sup>#</sup> |

Fishers exact test<sup>#</sup>; Kruskal-Wallis rank sum test<sup>‡</sup>.

### Successful development of antibodies in PLWH after two doses of BNT162b2 vaccination regardless of baseline CD4**^+^** T-cell counts

Serological responses against SARS-CoV-2 vaccination were assessed longitudinally in PLWH cohort by Elecsys assay before the first vaccination (T0), 1-month after the first dose (T1), and 3-months (T2) and 6-months (T3) after the 2nd dose. At T0, very low levels of anti-S titers were observed except for one PLWH participant showing very high titers (291 binding antibody units (BAU)/mL). To exclude any possibility of error, we repeated these experiments with anti-RBD assay that has a lower limit of quantification (Mesoscale Discovery, MSD, assay) that confirmed Elecsys data with strong correlation between the two assays (Pearson R=0.998; P<0.001). A significant rise in anti-RBD titers against the Wuhan-Hu-1 strain was observed at T1 in both PLWH groups, compared to T0 (P<0.01) (**Figure 1A**). While quantitative anti-S titers between PLWH-high and PLWH-low were not significantly different at T1, PLWH-low participants showed a significantly lower rate of seroconversion compared to PLWH-high (OR=7.0; P<0.001). However, by T2, anti-RBD titers significantly increased for both PLWH groups demonstrating “high” IgG titers according to the WHO International Reference for anti-SARS-CoV-2 immunoglobulins; and unexpectedly, patient H173 with CD4^+^ T-cell counts of 9 cells/mm^3^ also developed high anti-RBD titers (817 BAU/mL). By T3, these responses were maintained except for 2 PLWH-low participants including patient H173, where the titers dropped below the WHO-defined level of low protection (anti-RBD ≤45 BAU/mL; see Methods). However, no statistical difference in anti-S or anti-RBD titers was observed between controls and PLWH-high or PLWH-low (Figure 1A).

To study the impact of possible concurrent natural exposure of SARS-CoV-2 on the vaccine-driven anti-S and anti-RBD immunological responses, we also assessed anti-N IgG titers. At T0, 11/30 PLWH patients had measurable anti-N IgG titers with one participant showing highly elevated titers that also showed similarly elevated anti-S titers (**Figure 1C**). This participant belonged to the PLWH-low group treated with emtricitabine/tenofovir and dolutegravir and had HIV-RNA of 33 copies/mL with CD4^+^ T-cell counts of 166 cells/mL (average counts over 6-months of 171 cells/mm^3^). A very strong correlation was observed between anti-N and anti-RBD titers at all timepoints, including T0 (R=0.99; P<0.001) suggesting that some of our PLWH participants might already have been exposed to SARS-CoV-2 at T0. More importantly, these data suggest that PLWH under ART including PLWH with extremely low CD4^+^ T-cell counts, are fully competent to develop anti-S IgG response to BNT162b2 vaccine. We also demonstrate that serological responses are durable till 6 months after vaccination in PLWH, including most of the PLWH-low participants, and are fully comparable to serological responses sustained in healthy individuals.

### BNT162b2 vaccination-driven anti-SARS-CoV-2 antibodies in PLWH are broadly neutralizing

As CD4^+^ T-cells play an essential role in production of high-affinity neutralizing antibodies^22,23^, we first studied the quality of antibody response for Wuhan-Hu-1 by an ACE2 neutralization assay at T2 that showed the highest serological anti-S titers. The ACE2 neutralizing capacities for Wuhan-Hu-1 spike correlated significantly with the anti-S Wuhan-Hu-1 IgGs (Pearson’s R=0.831, P<0.001; Spearman ρ=0.848, P≤0.001), with only two PLWH-low participants showing lower than expected neutralization potential of their measured serological titers (**Figure 2A**).

**Figure 2.**
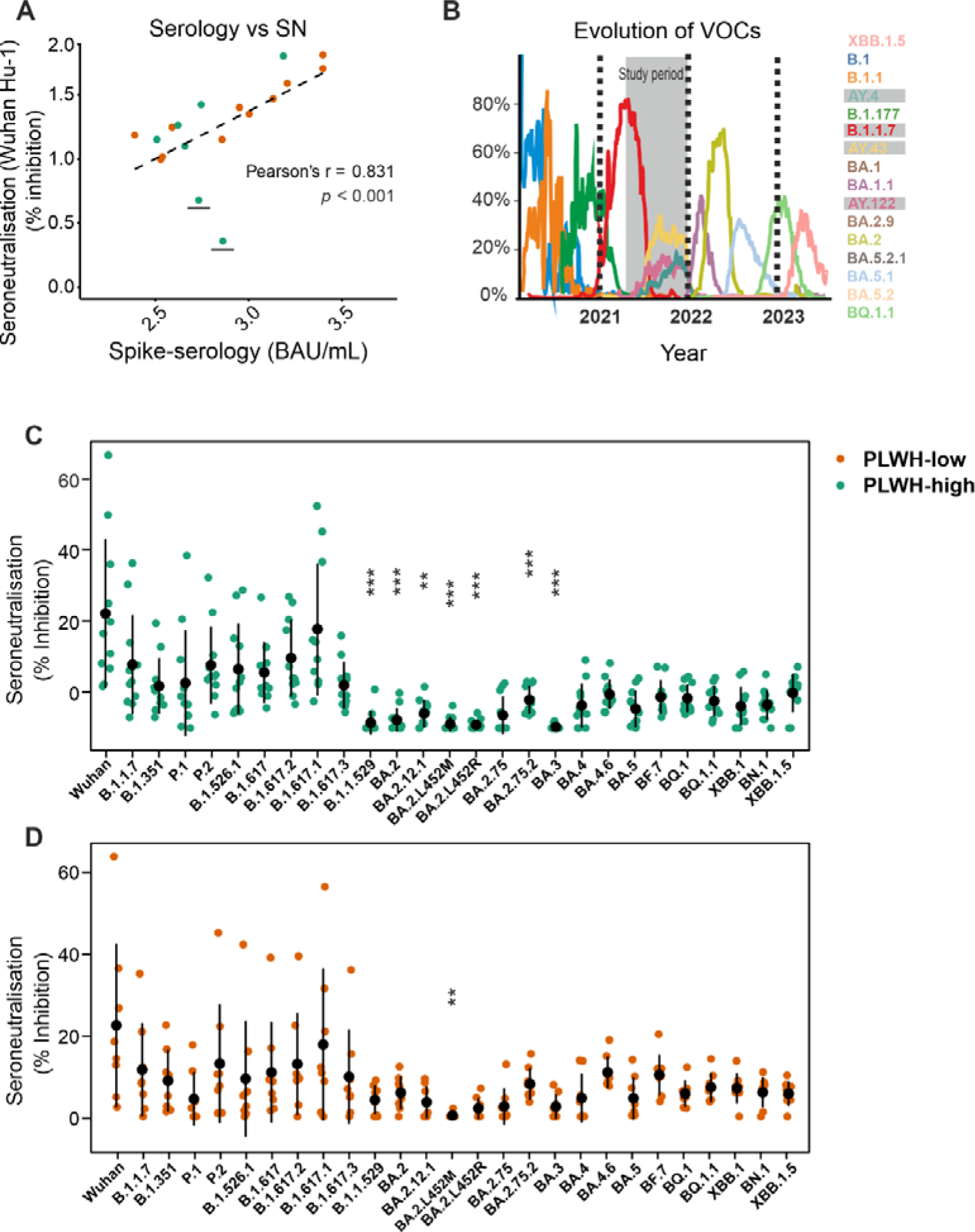
Anti-SARS-CoV-2 serological and seroneutralization responses in PLWH vaccinated with BNT162b2 vaccine. (A) Correlation between anti-RBD Serology and neutralization of Wuhan variant at 3 months after the second vaccination (T2). Two PLWH-low participants had high anti-RBD titers but neutralization potential was poor, marked with a black line. (B) Circulating VOCs in Italy during the study period. (C and D) Anti-Spike neutralization in PLWH-high and PLWH-low against different VOCs measured at T2. Dot plots indicate mean and whiskers indicate 1 standard deviation. **: p < 0.01. ***: p < 0.001.

While infection with different VOCs elicits variant-specific antibodies, prior mRNA vaccination in healthy populations has been found to imprint serological responses toward Wuhan-Hu-1 rather than variant antigens ^17,18^. As Alpha (B.1.1.7) was dominantly circulating in Italy during March-April 2021 at the time of vaccination, followed by the Delta (B.1.617) during July-August 2021, which was the follow-up period of our cohort, we studied the neutralization potential of sera from PLWH for these and other newer VOCs including XBB variants (**Figure 2B**). As expected, a decrease in neutralizing activity was found against each VOC from the early phases of the pandemic to currently circulating VOCs. The neutralization potential of anti-S IgGs was generally lower in PLWH-low than that of PLWH-high not only for Wuhan-Hu-1 (28% reduced) but for other VOCs. Sera from both PLWH-high and PLWH-low groups showed lower neutralizing activity against Omicron and its sub- variants when compared to Wuhan-Hu-1, although these differences were not significant for several variants such as BA.4.5 and XBB.1.5 (**Figure 2C, D**). These data suggest that while PLWH-high and PLWH-low can develop high titers of anti-S antibodies, these antibodies are also broadly neutralizing for ACE2 binding to different SARS-CoV-2 VOCs, although with different efficacies.

### T-cell responses are primed by subtle SARS-CoV-2 exposure that drives rapid antibody responses

Patients with HIV display an inversion in the ratio of CD4^+^ to CD8^+^ T-cells, which can be restored by ART to a large extent ^24^. We analysed CD4^+^ to CD8+ T-cell frequencies by flow cytometry over the entire study period and compared these to controls at T3. PLWH-high generally showed slightly increased CD4^+^ and CD8^+^ T-cells compared to PLWH-low at T0 and later timepoints, even if these differences were largely non-significant (**Figure 3, upper and middle panels**). However, compared to controls, CD4^+^ T-cells, expectedly, were lower in PLWH-low (P<0.035). Similarly, CD4:CD8 ratio for PLWH-low was also significantly lower than PLWH-high and controls (P<0.001, for both) (**Figure 3, lower panel**).

**Figure 3.**
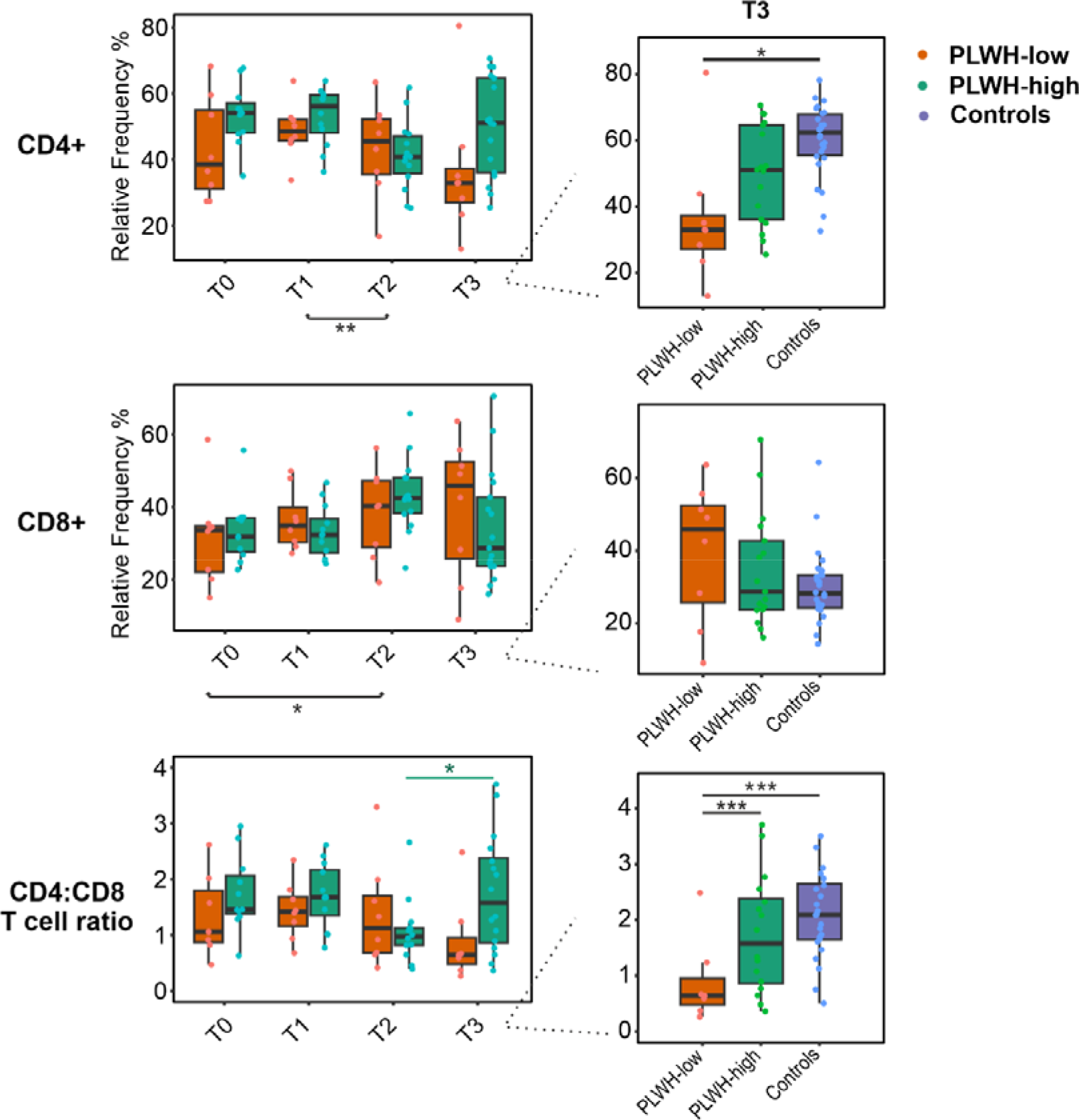
Evolution of CD4+, CD8+, and CD4:CD8 T-cells in PLWH and controls vaccinated with BNT162b2 vaccine. PLWH were studied from T0 till T3, while the controls were only studied at the last time point. Overall frequencies of CD4+ T cells of PLWH-high and PLWH-low were not significantly different during the study period, suggesting stable HIV disease status. Green statistical line represents the significance of pairwise comparisons overtime within PLWH-high group. Black significance lines under the plot represents significant groupwise comparisons overtime of PLWH group. Box plots indicate median (middle line), 25th and 75th percentile (box), and the 5th and 95th percentile (whiskers). All data points, including outliers, are displayed. PLWH, people living with HIV; T0: prior to the 1st dose vaccine; T1: prior to the 2nd dose; T2: 3 months after 2nd dose; T3: 6 months after 2nd dose. *: p < 0.05. **: p < 0.01. ***: p < 0.001.

Next, S-specific surface activation and cytokine expression markers in CD4^+^ and CD8^+^ T-cells were investigated in PLWH. S-specific CD4^+^ activation, measured as CD4^+^CD154^+^ cells, significantly increased in both PLWH groups from T0 to T3, even if S- and N-specific CD4^+^CD154^+^ were already present in some individuals at T0 in both PLWH groups (**Table 2**). As intrafamilial studies have shown that exposure to SARS-CoV-2 can lead to only cellular immune response without seroconversion^25^, we questioned whether this PLWH population had already been similarly primed towards SARS-CoV- 2 before vaccination. As priming leads to robust secondary immune response, we studied whether S-specific T-cell immunity at T0 correlated with development of anti-S IgG at T2 (**Supplementary** Figure 2). We show here a very strong correlation between anti-S IgG titers at T1 with S-stimulated CD4^+^ memory T-cells (CD4^+^CD45RO^+^; Pearson’s R=0.962, P<0.001) as well as S-stimulated intracellular cytokine (tumour necrosis factor (TNF)-α, interferon (IF)-γ and interleukin (IL)-2– expressing CD4^+^ or CD4^+^CD45RO^+^ memory T-cells (R≥0.965, P<0.001, for all) (Supplementary Figure 2), while the correlation with CD4^+^CD154^+^ cells was less strong (R=0.793, P<0.001). These data are not entirely surprizing as CD154 protein (CD40 ligand) while critical to the regulation of both humoral and cellular immune responses, the receptor is expressed only transiently^26^ and is less robust than memory or intracellular cytokine-expressing T-cells. A similar association of IgG titers was also observed with CD8^+^ cells but was not entirely independent as CD4^+^ and CD8^+^ T-cells also highly correlated for all activation markers (R≥0.96; P<0.001 for all) except for CD154, which is not expressed on CD8 cells. No correlation was observed between these T-cell activation markers and anti-S IgG titers measured at later timepoints likely because the titers plateaued after T1.

**Table 2.** Table highlighting changes in frequencies of CD4+ T-cell subsets following SARS-CoV-2 vaccination in PLWH.

| Characteristic Expressing (% [95% CI]) | Pre-vaccination N = 20 | V1M1 N = 20 | V2M3 N = 20 | V2M6 N = 20 | Characteristic Expressing (% [95% CI]) | Pre-vax vs V1M1 p-value | Pre-vax vs V2M3 p-value | Pre-vax vs V2M6 p-value | V1M1 vs V2M3 p-value | V1M1 vs V2M6 p-value | V2M3 vs V2M6 p-value |
| --- | --- | --- | --- | --- | --- | --- | --- | --- | --- | --- | --- |
| **Within CD4+ T cells** |  |  |  |  | **Within CD4+ T cells** |  |  |  |  |  |  |
| **Spike** |  |  |  |  | **Spike** |  |  |  |  |  |  |
| IFN-γ+ | 0.62 [0.30, 1.49] | 0.49 [0.29, 1.13] | 0.38 [0.22, 0.87] | 0.29 [0.10, 0.59] | IFN-γ+ | 1.00 | 1.00 | 1.00 | 1.00 | 0.80 | 1.00 |
| IL-2+ | 1.71 [0.76, 4.05] | 2.47 [1.15, 3.88] | 1.67 [0.80, 2.96] | 2.58 [0.45, 5.46] | IL-2+ | 1.00 | 1.00 | 1.00 | 1.00 | 1.00 | 1.00 |
| TNF-α+ | 1.67 [0.73, 3.43] | 1.65 [0.93, 3.04] | 1.18 [0.78, 1.89] | 0.03 [0.01, 1.59] | TNF-α+ | 1.00 | 1.00 | 1.00 | 1.00 | 1.00 | 1.00 |
| CD154+ | 1.52 [1.16, 3.22] | 2.07 [1.54, 4.21] | 1.02 [0.37, 3.16] | 8.43 [4.54, 13.20] | CD154+ | 1.00 | 1.00 | **0.01** | 0.63 | **0.01** | **0.00** |
| CD45RO+ | 26.05 [19.08, 34.08] | 23.80 [19.73, 27.60] | 33.90 [28.78, 36.33] | 25.50 [19.30, 33.50] | CD45RO+ | 1.00 | 0.80 | 1.00 | **0.01** | 1.00 | 1.00 |
| **Within CD8+ T cells** |  |  |  |  | **Within CD8+ T cells** |  |  |  |  |  |  |
| IFN-γ+ | 2.24 [1.21, 4.73] | 2.25 [0.82, 4.82] | 1.41 [0.37, 5.01] | 0.00 [0.00, 0.40] | IFN-γ+ | 1.00 | 1.00 | **0.02** | 1.00 | 0.19 | 0.32 |
| IL-2+ | 1.64 [1.23, 2.85] | 2.30 [1.27, 3.00] | 1.75 [0.56, 3.05] | 0.08 [0.01, 1.22] | IL-2+ | 1.00 | 1.00 | **0.00** | 1.00 | **0.04** | 0.24 |
| TNF-α+ | 3.97 [2.85, 9.12] | 4.54 [1.99, 8.72] | 3.93 [1.53, 10.55] | 0.02 [0.00, 0.06] | TNF-α+ | 1.00 | 1.00 | **0.01** | 1.00 | 0.08 | 0.12 |
| CD45RO+ | 8.68 [6.51, 10.78] | 10.30 [7.15, 11.73] | 10.20 [9.04, 15.65] | 10.60 [4.20, 13.20] | CD45RO+ | 1.00 | 0.24 | 1.00 | 1.00 | 1.00 | 1.00 |
| **Within CD4+ T cells** |  |  |  |  | **Within CD4+ T cells** |  |  |  |  |  |  |
| **Nucleocapsid** |  |  |  |  | **Nucleocapsid** |  |  |  |  |  |  |
| IFN-γ+ | 0.85 [0.37, 1.77] | 0.26 [0.11, 1.13] | 0.11 [0.05, 0.25] | 0.03 [0.02, 0.04] | IFN-γ+ | 1.00 | 0.06 | **0.00** | 0.46 | **0.00** | **0.03** |
| IL-2+ | 2.13 [1.61, 4.24] | 1.85 [1.08, 4.64] | 1.23 [0.34, 2.54] | 0.09 [0.04, 0.84] | IL-2+ | 1.00 | 1.00 | 1.00 | 0.32 | 0.92 | 1.00 |
| TNF-α+ | 2.22 [1.15, 4.54] | 0.69 [0.42, 3.06] | 0.57 [0.25, 0.88] | 0.07 [0.00, 1.13] | TNF-α+ | 0.78 | **0.01** | 0.38 | 0.24 | 0.52 | 1.00 |
| CD154+ | 1.67 [1.34, 3.67] | 1.80 [0.88, 4.72] | 0.63 [0.25, 2.29] | 4.22 [2.16, 9.47] | CD154+ | 1.00 | 0.54 | **0.02** | 0.54 | 0.12 | **0.01** |
| CD45RO+ | 25.10 [20.00, 33.05] | 22.95 [19.10, 31.80] | 34.65 [29.05, 40.53] | 21.00 [14.30, 32.80] | CD45RO+ | 1.00 | 0.09 | 1.00 | 0.08 | 1.00 | 0.08 |
| **Within CD8+ T cells** |  |  |  |  | **Within CD8+ T cells** |  |  |  |  |  |  |
| IFN-γ+ | 4.43 [2.51, 12.05] | 2.13 [0.38, 5.89] | 0.28 [0.11, 0.80] | 0.01 [0.00, 0.06] | IFN-γ+ | **0.01** | **0.02** | **0.00** | 0.07 | **0.00** | 0.18 |
| IL-2+ | 1.94 [1.49, 3.51] | 1.61 [0.90, 3.53] | 0.78 [0.35, 1.94] | 0.02 [0.00, 0.36] | IL-2+ | 1.00 | 0.16 | 0.06 | 0.50 | **0.00** | 0.86 |
| TNF-α+ | 8.83 [4.17, 17.08] | 3.99 [0.94, 8.21] | 1.00 [0.50, 2.15] | 0.09 [0.00, 0.67] | TNF-α+ | **0.01** | **0.00** | **0.00** | 0.22 | **0.00** | 0.97 |
| CD45RO+ | 8.81 [7.53, 12.68] | 9.68 [7.66, 11.10] | 10.65 [8.59, 18.75] | 10.10 [4.42, 15.20] | CD45RO+ | 1.00 | 0.63 | 1.00 | 0.40 | 1.00 | 1.00 |

**\*\*Wilcoxon pairwise rank sum test**
Results are presented as median (IQR) of the percentage (frequencies) within total live CD4+ T-cells (upper panel) or CD8+ T cells (lower panel) within spike- and nucleocapsid- stimulated T-cells. Statistics used - Wilcoxon pairwise rank sum test. Study visits - T0: pre-vaccination baseline, T1: 1 month post 1st dose, T2: 3 months post 2nd dose, T3: 6 months post 2nd dose.

Similar to serological data, the S- and N- specific memory T-cell marker remained stable throughout the study period and did not significantly differ between the PLWH-high and -low groups (Table 2). Moreover, no significant difference between PLWH and controls was observed for S- and N- specific CD4^+^CD45RO^+^ or CD8^+^CD45RO^+^ cells. These data suggest that despite having a reduced CD4^+^ T-cells and CD4:CD8 ratio, PLWH including PLWH-low participants develop and sustain a robust SARS-CoV- 2–specific T-cell and memory T-cell response.

### PLWH show preferential expression of IL-2, but not IFN- **γ**, in SARS-CoV-2-specific T-cell activation

Polyfunctionality of effector T-cells at the single cell level has been shown as an important parameter to predict the quality of T-cell responses and immunological control of infectious diseases including SARS-CoV-2 ^5,27,28^. We therefore addressed intracellular expressions of TNF-α, IFN-γ, and, IL-2 in CD4^+^ and CD8^+^ T-cells in PLWH population and compared them to controls (**Figure 4A, B**). While the cytokine expression of S-specific IL-2, TNF-α, and IFN-γ in CD4^+^ T-cells did not significantly change during the study period, a declining trend was observed by T3 (Table 2), as observed earlier^29^. CD4^+^ or CD8^+^ T-cells co-expressing IFN-γ^+^TNF-α^+^IL-2^+^ were much more frequent in controls, however, did not reach statistical significance with either of the PLWH groups. Similarly, co-production of two cytokines was also rare in PLWH with a noticeable difference for IFN-γ^+^IL-2^+^ cells for both PLWH-high and PLWH-low groups, although again these differences did not reach statistical significance. The majority of analysed T-cells in all study groups expressed only one of the three studied cytokines. Remarkably, the majority of CD4^+^ T-cells in controls produced a significantly higher amount of IFN-γ, which was significantly different between control and PLWH-low (P<0.05). On the other hand, PLWH-low and PLWH-high demonstrated a significantly higher frequency of IL-2- expressing CD4^+^ compared to controls (6 to 56-fold, respectively; P<0.001, for both). PLWH-low also showed 7-fold significantly higher frequencies of CD4^+^IL2^+^ cells compared to PLWH-high (P=0.014). Remarkably, the patient with the lowest absolute CD4^+^ T-cell counts (patient H173) had the highest proportion of S-specific CD4^+^IL-2 expression and S-specific CD4^+^CD45RO^+^ memory T-cells. Moreover, CD4 counts in PLWH and controls showed a strong negative correlation with both IL-2-expressing CD4^+^ T-cells and CD4^+^ memory T-cells (Spearman Rho: -0.85 and -0.80, respectively; P<0.001). These data suggest that the subset distribution in PLWH of IFN-γ, TNF-α, and IL-2 was dominated by a higher proportion of monofunctional T-cells and lower proportions of bi-functional or triple-functional cells. More importantly, these data suggest that PLWH preferentially respond with utilizing IL-2 in their SARS-CoV-2-specific T-cell effector functions.

**Figure 4.**
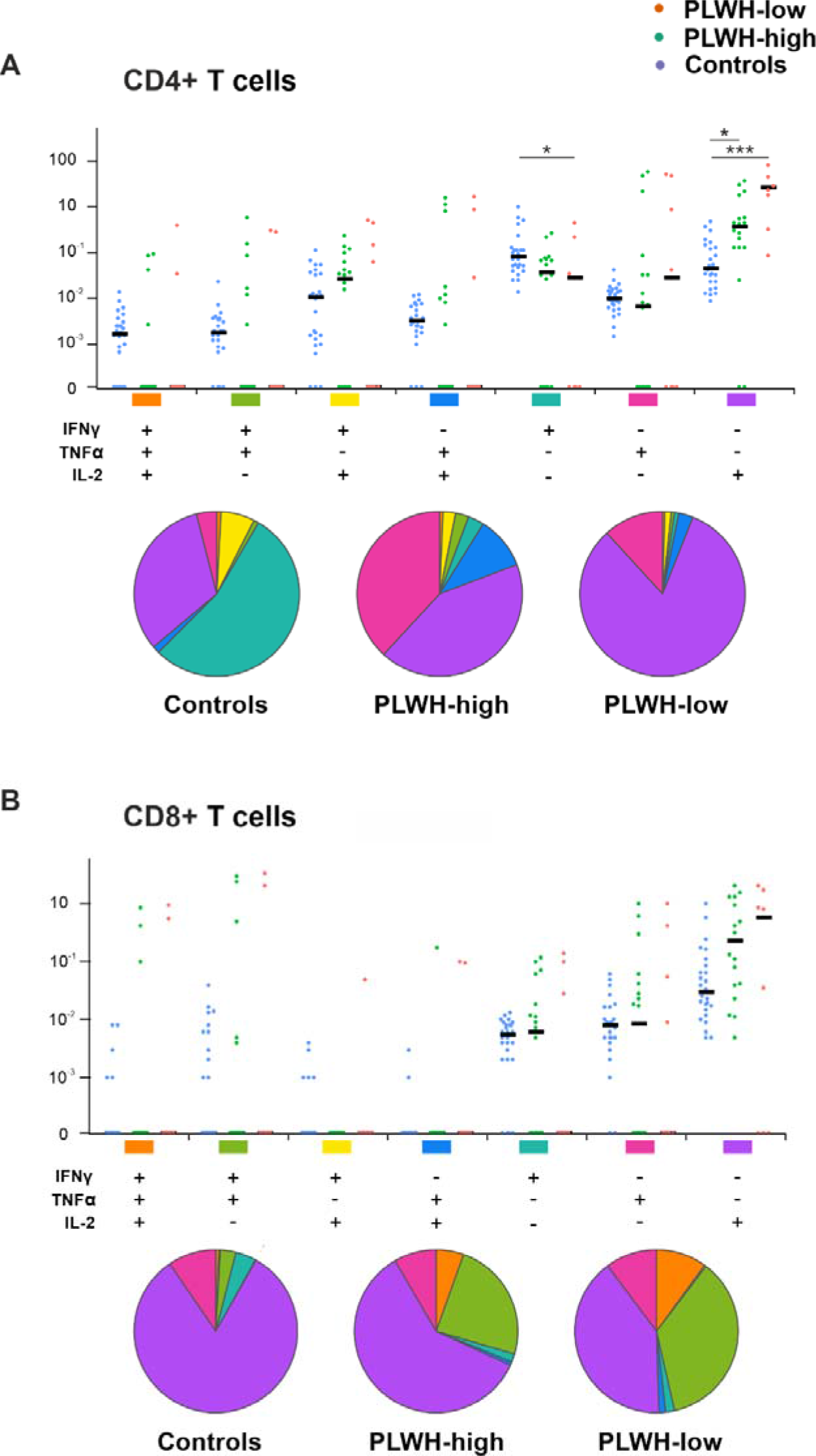
Impact of BNT162b2 vaccination on SARS-CoV-2 spike specific polyfunctional cytokine expression profiles in CD4^+^ and CD8^+^ T-cells. Profiles of individual and simultaneous cytokine expression in PLWH and HIV-negative control participants vaccinated with BNT162b2 vaccines analysed at 6 months after the second vaccination (T3) after stimulating with SARS-CoV-2- specific S- peptides. The size of each pie segment relates to the frequency of a monofunctional, bifunctional and triple-functional response. Statistics was performed using SPICE software for polyfunctionality assessment (see Methods).

## Discussion

In this study, we show that ART-treated PLWH with optimal virological suppression vaccinated with BNT162b2 mRNA vaccine despite demonstrating lower CD4^+^ T-cell counts and incompletely restored CD4:CD8 ratio, generate robust SARS-CoV-2–specific CD4^+^ and CD8^+^ T-cell responses that are sustained till 6 months after the second dose. Except for two PLWH-low participants (both with CD4^+^ T-cell counts of <200 cells/mm^3^), the serological response in the PLWH was remarkably similar to that noted for healthy controls. We also show that CD4^+^ T-cells completely corroborate with the serological data except for the pre-vaccination timepoint where presence of S- and N-specific CD4^+^ T-cells was evidenced without notable serological titers. Although low-level T-cell epitope cross-recognition with other coronaviruses has been reported^30^, we showed that these T-cell responses were specific as S-activated primed CD4^+^ T-cells present at pre-vaccination timepoint correlated highly with development of anti-S IgG titers one month after the first vaccination dose. T-cell immune responses without seroconversion have been reported in intrafamilial SARS-CoV-2 exposure involving healthy individuals^25^, however, silent exposure to SARS-CoV-2 in PLWH including individuals with CD4^+^ counts < 200 cells/mm^3^ has not been previously reported. Notably, while worse outcomes should be expected in HIV-positive patients compared to the general population, no evidence so far of any increased rates of COVID-19 disease compared to general population has been reported^31,32^. Paradoxically, COVID-19 patients with more severe HIV-related immunosuppression have been reported to experience only a mild-to-moderate disease^5,33^. In our study, one PLWH-low participant showed evidence of heavy viral exposure before vaccination, reflected by the highest high S- and N- titers of all studied participants, yet did not report being infected. These data are in line with vaccination studies where HIV patients on ART have been shown to be competent enough to develop and sustain vaccine response to SARS-CoV-2^9,11,12^.

While the mechanisms remain elusive, several mutually nonexclusive factors could be involved. Firstly, the number of CD4^+^ T-cells for any specific immunity reaction is kept optimal by limiting the proliferation of effector T-cells, such as through CTLA-4-dependent mechanisms, or having T effector cells undergo apoptosis after generating the expected response. This occurs because of a decline in antigen levels thereby depriving the effector cells of the required level of antigen and cytokine stimulation to survive. While that is a very general phenomenon in healthy individuals, in HIV patients, an enhanced propensity to apoptosis has been observed that is not completely reversed by ART^34^. Secondly, it is clear that HIV infection does not deplete SARS-CoV-2-specific CD4^+^ T-cells. This is of relevance as HIV preferentially affects HIV-specific CD4^+^ T-cells, however, clonal deletion of several non-HIV antigen-specific cells is also reported^35^. Moreover, while CD4^+^ memory T-cells are a major reservoir for HIV in ART-treated individuals, depletion of SARS-CoV-2-specific CD4^+^ memory T-cells also does not seem to occur as these cells persisted in high numbers through the study period including 6 months after the second dose. Thirdly, appropriate stimulation of T-cells also supports data that incompletely eradicated virus in ART-treated PLWH participants leads to persistent immune activation with increased production of immune-stimulatory cytokines that activate T-cells^36^. Thus, it is possible that these proinflammatory aspects of latent HIV infection could facilitate the activation and proliferation of SARS-CoV-2-specific CD4^+^ T-cells^5,33^.

Lastly, we show that PLWH on ART utilizes IL-2 as one of the chief effector cytokines specifically for CD4^+^ T-cells, at least against SARS-CoV-2 antigens. IL-2 is secreted primarily from activated CD4^+^ T-cells and mediates several biological processes that promote immune system function. Several *in vitro* studies have also demonstrated the effects of IL-2 on several parameters of immune function in HIV-infected individuals^37^. For instance, IL-2 induces T-cell proliferation, promotes the release of secondary cytokines, and is a critical determinant of the fate decisions of antigen receptor–activated T-cells^38,39^. It also activates B-cells thereby promoting antibody synthesis^40^ and decreases the rate of lymphocyte apoptosis^41^. With such important roles of IL-2 in HIV, not surprisingly, prior to the advent of ART therapy, human recombinant IL-2 (rIL-2) was evaluated in clinical trials in HIV-infected patients to promote immune system recovery, however, it was associated with increased toxicity^42^. Given our data of IL-2-expressing CD4^+^ T-cells associated with a robust immune response in PLWH, it would be tempting to suggest that IL-2-based compounds could be re-considered as an adjuvant therapy in patients not fully responsive to ART. To support this premise, despite earlier failure of rIL- 2 in cancer and autoimmune diseases again due to toxicities, modified IL-2-based adjuvants and targeted therapies are currently being tested in early-stage to phase 3 clinical trials for these disorders^43^.

As low viremia and persistently low CD4^+^ T-cell counts observed in PLWH-low participants in our cohort could potentially affect the epitope recognition capacity and alter the quality of T-cell responses, we also studied ACE2 neutralization for 27 different SARS-CoV-2 variants and show an optimal quality of CD4^+^ T-cell responses with expected ACE2 neutralizing capacities. Importantly, our data shows that PLWH have also a lower chance of imprinting, a mechanism that ensures to recall existing memory cells rather than stimulating a *de novo* response when the immune system encounters a new but closely related antigen^44,45^. This is because PLWH participants exposed to SARS-CoV-2 before receiving vaccination were likely exposed to SARS-CoV-2 B.1.1.7 or B.1.617 VOCs but developed a robust response to BNT162b2 mRNA vaccine encoding Wuhan Hu-1 spike protein. These data suggest that combination vaccines such as bivalent boosters covering newer Omicron VOCs would continue to protect PLWH from SARS-CoV-2 with high efficacy with little interference of imprinting.

As limitations, we have studied only a limited number of PLWH participants necessitating the need for larger studies to assess the role of IL-2 in T-cell activation in PLWH. The lack of longitudinal data for controls prevented us from being able to compare immune dynamics in PLWH with a HIV-negative population. Additionally, T-cell responses against relevant S-specific VOCs along with long-term follow-up should be undertaken to study whether immune memory is better sustained in controls to guide the frequency of boosting in PLWH.

Despite these limitations, utilizing a comprehensive longitudinal data set on humoral and cellular immune responses in PLWH and age- and vaccination-matched controls we show that PLWH individuals treated with ART including those with CD4^+^ T-cell counts <200 cells/mm^2^ have adaptive cell immunity that is sufficiently competent to generate humoral and cellular responses against BNT162b2 mRNA. We also show that PLWH preferentially utilizes IL-2 for their effector functions. Whether this is also a general mechanism that PLWH utilizes to make their deficient CD4^+^ T-cell more efficient remains to be studied.

## Material and Methods

### Study population and design

Participants were recruited from the Infectious Diseases Section of the University Hospital of Verona as a part of the prospective, multicentric ORCHESTRA study ^46^. A total of 56 participants were enrolled, of which 30 PLWH and 26 healthcare worker controls matched for age and vaccination timepoint. All participants provided informed, written consent before study participation. The study was approved by the University Hospital Verona Ethics Board and conducted in accordance with the Declaration of Helsinki. Serum, plasma, and peripheral blood mononuclear cell (PBMC) samples were collected between April and December 2021. For each PLWH, sample collection occurred at multiple timepoints after receiving the BNT162b2 vaccine: (i) T0, just prior to the first dose; (ii) T1, just prior to the second dose; (iii) T2, three months after the second dose ± 4 days; and (iv) T3, six months after the second dose ± 4 days. An overview of sample collection is available in **Supplementary** Figure 1.

### Serology

Anti-RBD SARS-CoV-2 IgG titers were measured in serum samples at University Hospital of Verona using the Elecsys Anti-SARS-CoV-2 Spike (Roche Diagnostics, Basel, Switzerland) immunoassay). Serological analyses was also performed in duplicate on anti-N, anti-S, and anti-RBD SARS-CoV-2 IgG titers in plasma samples at Molecular Pathology-CBH Laboratory of University of Antwerp. IgG titers were measured in plasma samples using V-PLEX Panel 2, 32, and 34 (IgG) Kit (Meso Scale Discovery, MSD, Maryland, USA) according to the manufacturer’s instructions ^47^.

### ACE2 neutralization

ACE2 neutralization was measured in duplicate in plasma samples using V-PLEX Panel 13, Panel 27, and Panel 32 Kits (ACE2) (MSD) according to the manufacturer’s instructions. The following SARS-CoV-2 variants were measured: Wu-Hu-1 (Wuhan), B.1.1.7 (Alpha), B.1.351 (Beta), P.1 (Gamma), B.1.617.3 (Delta), and Omicron with sub-variants - BA.1, BA.2, BA.2+L452M, BA.2+L452R, BA.2.75, BA.2.75.2, BA.3, BA.4, BA.4.6, BA.5, BF.7, BQ.1, BQ.1.1, BN.1, XBB.1, XBB1.5.

### SARS-CoV-2 specific T-cell responses

PBMCs were isolated and frozen at -80°C in foetal bovine serum (FBS)/10% dimethyl sulfoxide (DMSO) at the collection site before being transported for analyses. Both PLWH and control individuals were collected at the same centre with harmonized protocols. On the day of analysis, PBMCs were thawed and rested overnight. The cells were counted and all aliquots of PBMC were pooled together and selected for processing. Activation induced marker (AIM) assay was performed, optimised from earlier protocols ^48^. For stimulation, spike glycoprotein (“Protein S”; Prot_S, GenBank MN908947.3, Protein QHD43416.1) and nucleocapsid phosphoprotein (“NCAP”; GenBank MN908947.3, Protein QHD43423.2) of SARS-CoV-2 (JPT peptide technologies, Berlin, Germany) were utilized. Further details can be found in supplemental methods.

### T-cell staining and flow cytometry

Peptide-stimulated cells and controls were stained with a fixable viability dye for 15 minutes in the dark at room temperature. Cells were washed with cell staining buffer (PBS 1% bovine serum albumin, 2mM EDTA) unless stated otherwise. Cells were then stained with surface antibody mixture, see **Supplemental Table 1**, for 15 minutes at room temperature. Afterwards, cells were washed with cell staining buffer and fixed/permeabilized for 20 minutes in the dark at room temperature. Then, cells were washed with permeabilization buffer and stained with antibodies directed towards intracellular cytokines (Supplemental Table 1) for 15 minutes in the dark at room temperature. Finally, cells were washed in cell staining buffer. Flow cytometry (FC) was performed on NovoCyte Quanteon 4025 flow cytometer (Agilent, CA, USA), and FC data analysed using FlowJo– v10.8.1 (BD Biosciences, CA, USA).

### Data analysis and statistics

All data were collected using Microsoft Excel. Visualization and statistical analysis of all data was done in Studio (R version 4.2.2). Titers of binding RBD-, S and N-specific IgG as well as titers of neutralizing Abs were visualized in dot plots indicating average and ±1 standard deviation (SD). T-cell responses over time and compared to controls were visualized in boxplots unless mentioned otherwise in the legend. After assessing normality, serological data was analysed using parametric statistical methods and cellular data was analysed using non-parametric statistical methods. Post hoc p-value correction was conducted using Bonferroni’s correction method for all analyses. Pearson’s and Spearman correlation were utilized to assess the relationship between serology and seroneutralization data, and all other T-cell markers. Distribution differences of the different cytokine combinations between the groups were performed using SPICE (see supplemental methods).

### Study approval

Ethical approval for this study was obtained from the University Hospital Verona Ethics Board (protocol number: 19293) and conducted in accordance with the Declaration of Helsinki. No study activities took place prior to collection of informed consent.

## Data availability

Data supporting the findings of this study are available within Supplementary Information files. All other data generated in this study are available from the corresponding author upon request.

## Author Contribution

Overall study supervision and conceptualization: SKS; Study design ER, AG, MM, SK-S; Clinical data and sample collection: ER, MB, CS, MM, AS, AA, DP, MGLM, SP, ET; Serological analysis: AG, AK, AH; PBMC isolations: ER, CS, GS, AS; PBMC analysis: AG, AK, SW, VVA; Statistical analysis: AG, SKS, MM; Data interpretation: SKS, AG, ER, GV; Data visualization: AG; Manuscript writing: SKS, AG; Manuscript review: AG, ER, AH, SMK, GV, SP, ET, SKS. All authors read, gave input, and approved the final manuscript. The authors thank Lorenzo Maria Canzani (University of Verona) for critical reading.

