## Supplementary Information for "IL-2-mediated CD4 T-cell activation correlates highly with effective serological and T-cell responses to SARS-CoV-2 vaccination in people living with HIV (PLWH)"

* Shared first authorship

† Shared senior authorship

**#Correspondence**: Samir Kumar-Singh, Faculty of Medicine & Health Sciences, Vaccine & Infectious Disease Institute, University of Antwerp – CDE – T1.32, Universiteitsplein 1, 2610 Antwerp, Belgium, +32 3 265 3329,

**Funding**: This work is supported by the ORCHESTRA project, which has received funding from the European Union’s Horizon 2020 research and innovation program under grant agreement No 101016167.

**Conflicts of interest**: None of the authors declare any conflict of interest related to this paper.

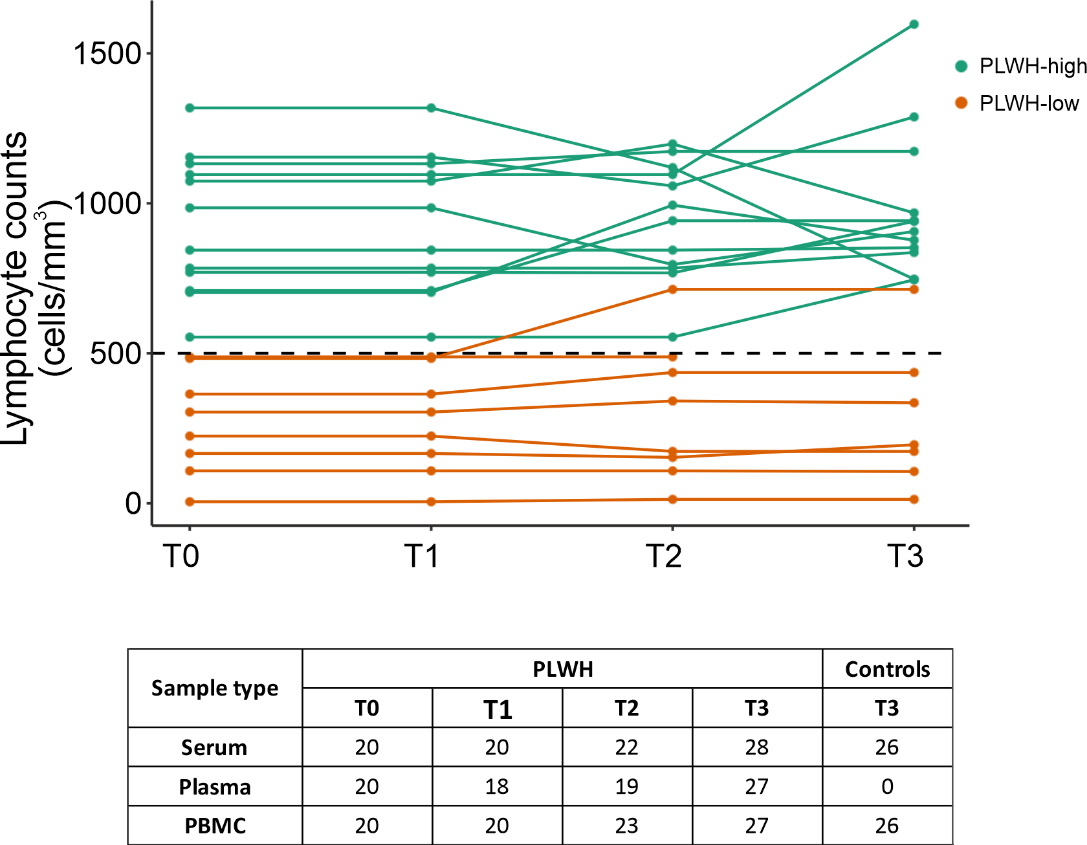

**Supplementary Figure 1. Study period overview and summary of samples collected per timepoint.** Circulating variants and VOCs have been demonstrated in Italy during the study period. The clinical CD4 counts demonstrating HIV disease progression are demonstrated during the sample collection period. Serum, plasma, and PBMC samples were collected at T0, T1, T2, T3 for PLWH. At T3, only serum and PBMC samples were collected for HCW controls. PBMC: peripheral blood mononuclear cell; PLWH: people living with HIV; HCW: healthcare worker. T0: Pre-vaccination; T1: 1 month after the first vaccination; T2: 3 months after the second vaccination; T3: 6 months after the second vaccination. Figure designed with Bio Render (Toronto, Ontario, Canada).

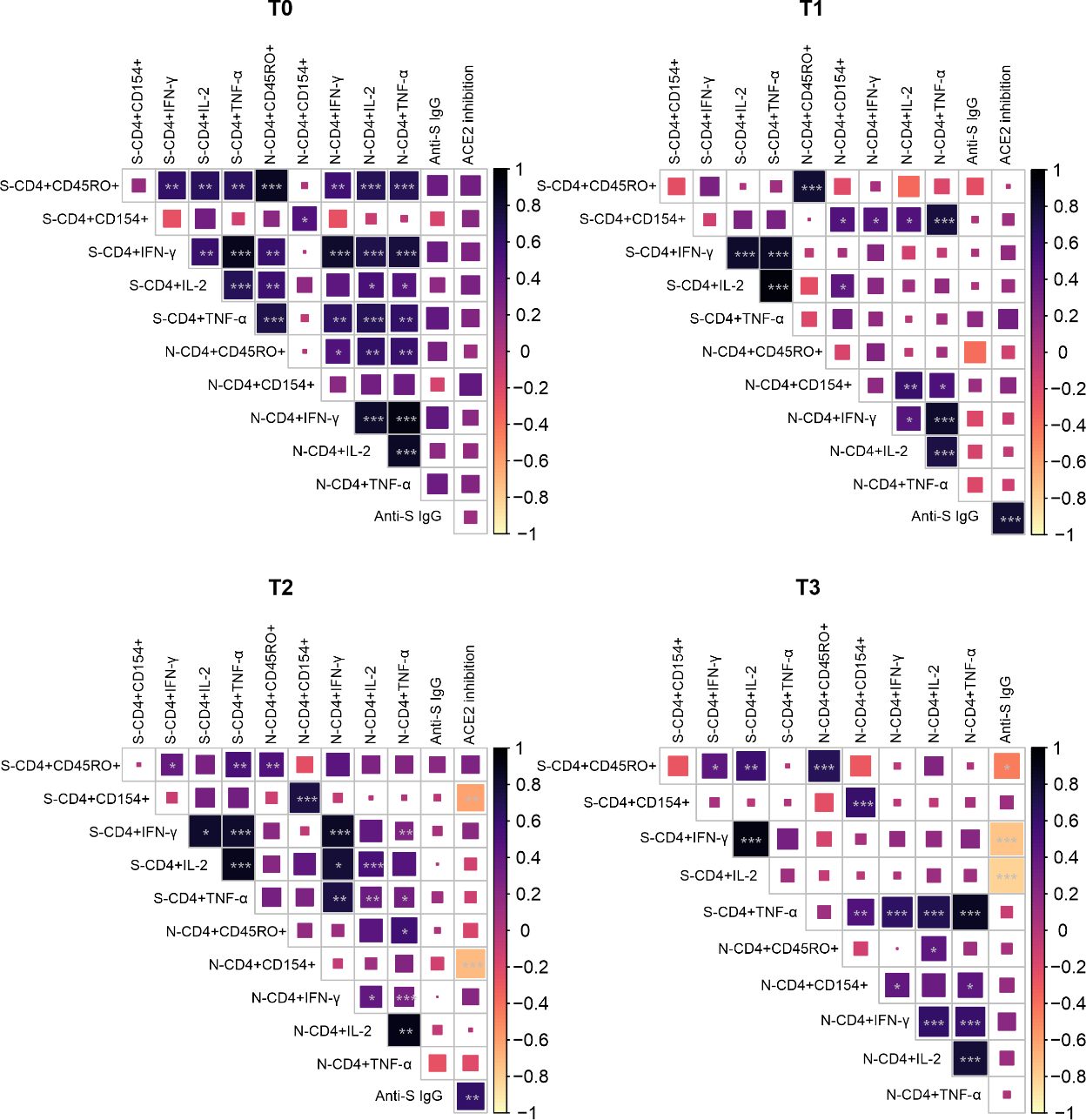

**Supplementary Figure 2. Correlation matrix depicting correlation between the various T cell and humoral parameters investigated.** IgG antibody levels to SARS-CoV-2 and percentage ACE2 inhibition are log_10_ transformed. Dark blue indicates good correlation and light yellow indicates poor corelation. Pearson’s correlations were considered significant when p <0.05. *: p < 0.05. **: p < 0.01. ***: p < 0.001. RF: Relative frequency, S: SARS-CoV-2 spike specific peptides, N: SARS-CoV-2 nucleocapsid-specific peptides. T0: pre-vaccination baseline, T1: 1 month post 1st dose, T2: 3 months post 2nd dose, T3: 6 months post 2nd dose.

**Material and** **Methods**

**Study population and design**

A total of 56 participants were enrolled, of which 30 PLWH and 26 healthcare worker (HNC) controls matched for age and vaccination timepoint. All participants provided informed, written consent before study participation. The study was approved by the University Hospital Verona Ethics Board and conducted in accordance with the Declaration of Helsinki. As for data collection, clinical data along with serum, plasma, and peripheral blood mononuclear cell (PBMC) samples were collected. Pairwise longitudinal data was available for 20 PLWH – PLWH-high (≥500/mm^3^; n=12) and PLWH-low (<500 cells/mm3; n=8). BD FACS Presto system was utilized to have CD4^+^ T-cell counts as a part of laboratory workup. The system is designed to provide absolute and percentage results of CD4^+^ T-lymphocytes. All samples were collected between April and December 2021. For each PLWH enrolled, sample collection occurred at multiple timepoints: : (i) T0, just prior to the first mRNA vaccine dose; (ii) T1, just prior to the second mRNA vaccine dose; (iii) T2, three months after the second mRNA vaccine dose ± 4 days; and (iv) T3, six months after the second mRNA vaccine dose ± 4 days. An overview of sample collection is available in **Supplementary Figure 1**.

**Serology**

Blood was collected in 10 mL BD Vacutainer serum tubes (BD Biosciences, NJ, USA). Anti-RBD SARS-CoV-2 IgG titres were measured in serum samples at University Hospital of Verona within 3h of blood collection. The Elecsys Anti-SARS-CoV-2 Spike (Roche Diagnostics, Basel, Switzerland) immunoassay was utilized and data was reported in WHO-recommended binding antibody units (BAU/mL). Serological analyses was also performed in duplicate on anti-N, anti-S, and anti-RBD SARS-CoV-2 IgG titres in plasma samples at Molecular Pathology-CBH Laboratory of University of Antwerp. IgG titers were measured in plasma samples using V-PLEX Panel 2, 32, and 34 (IgG) Kit (Meso Scale Discovery, MSD, Maryland, USA) according to the manufacturer’s instructions.

**ACE2 neutralization**

Blood was collected in 10 mL plasma tubes (EDTA plasma, BD Vacutainer PPT, BD Biosciences). ACE2 neutralization was measured in duplicate in plasma samples using V-PLEX Panel 13, Panel 27, and Panel 32 Kits (ACE2) (MSD) according to the manufacturer’s instructions. IgGs capable of blocking the binding of ACE2 to the following SARS-CoV-2 variants were measured: Wu-Hu-1 (Wuhan), B.1.1.7 (Alpha), B.1.351 (Beta), P.1 (Gamma), B.1.617.3 (Delta), and Omicron with sub-variants - BA.1, BA.2, BA.2+L452M, BA.2+L452R, BA.2.75, BA.2.75.2, BA.3, BA.4, BA.4.6, BA.5, BF.7, BQ.1, BQ.1.1, BN.1, XBB.1, XBB1.5.

**SARS-CoV-2 specific T-cell responses**

PBMCs were isolated using Cell Preparation Tubes, CPTs and frozen at -80°C in foetal bovine serum (FBS)/10% dimethyl sulfoxide (DMSO) at the clinical collection site before being transported on dry ice with temperature in-transport monitoring for analyses. Both PLWH and HNC individuals were collected at the same centre with harmonized protocols. On the day of analysis, PBMCs were thawed and rested overnight in RPMI 1640 medium (Gibco, Thermofisher Scientific, MA, USA) supplemented with 5% heat-inactivated AB serum (Sigma Aldrich), 100 U/mL penicillin (Biochrom, Berlin, Germany) and 0.1 mg/mL streptomycin (Biochrom). The cells were counted and all aliquots of PBMC were pooled together and selected for processing. Activation induced marker (AIM) assay was performed, optimised from earlier protocols ^53^. For stimulation, 2 pools of lyophilized peptides were utilized. The first pool was of 15-mer sequences with 11 amino acids (aa) overlap, covering the immunodominant sequence domains of the spike glycoprotein (“Protein S”; Prot_S, GenBank MN908947.3, Protein QHD43416.1) of SARS-CoV-2 (Miltenyi Biotech, Leiden, Netherlands). The second pool was a 102-peptide sequence of the nucleocapsid phosphoprotein (“NCAP”; GenBank MN908947.3, Protein QHD43423.2) of SARS-CoV-2 (JPT peptide technologies, Berlin, Germany). Both peptide pools were utilized at 1 μg/mL. Unstimulated negative controls were performed with equal volumes of sterile water and 10% DMSO. Cell activation cocktail, composed of PMA (phorbol-12-myristate 13- acetate) and ionomycin (Bio Legend, Amsterdam, Netherlands) in DMSO was used as positive controls. Incubation of PMBCs was performed at 37 °C, 5% CO2 for 2 hours after which 2 μg/mL brefeldin A (BioLegend, Amsterdam, Netherlands) was added and further incubation for 4 hours at 37°C and 5% CO2 before being collected for flow cytometric analysis.

**T-cell staining and flow cytometry**

Peptide-stimulated cells (as well as positive and negative controls) were stained with Zombie aqua fixable viability dye (BioLegend) for 15 minutes in the dark at room temperature. Cells were washed with cell staining buffer (PBS 1% bovine serum albumin, 2mM EDTA) unless stated otherwise. Cells were stained with surface antibody mixture including anti-CD3-APC Fire750, anti-CD4-FITC, anti-CD8a-BV570, anti-CD154-APC and anti-CD45RO-PerCP/Cyanine5.5 (BioLegend) for 15 minutes at room temperature. Afterwards, cells were washed with cell staining buffer and fixed/permeabilized using inside stain kit following manufacturer’s instructions (Inside stain kit, Miltenyi Biotec) for 20 minutes in the dark at room temperature. Then, cells were washed with permeabilization buffer and stained with antibodies directed towards intracellular cytokines (anti-IFNγ-PE, anti-TNFα-PE/Cyanine7 and anti-IL-2-BV421, BioLegend) for 15 minutes in the dark at room temperature. Finally, cells were washed in cell staining buffer. Flow cytometry (FC) was performed on NovoCyte Quanteon 4025 flow cytometer (Agilent, CA, USA), and FC data analysed using FlowJo v10.8.1 (BD Biosciences, CA, USA).

**Supplemental Table 1.** List of surface and intracellular staining antibodies.

| **Antibody** | **Clone** |
| --- | --- |
| **Surface Antibodies** | |
| APC anti-human CD154 | 24-31 |
| APC/Fire 750 anti-human CD3 | SK7 |
| PerCP/Cyanine5.5 anti-human CD45RO | UCHL1 |
| Brilliant Violet 570™ anti-human CD8a | RPA-T8 |
| FITC anti-human CD4 | RPA-T4 |
| **Intracellular antibodies** | |
| PE anti-human IFN-γ | 4S.B3 |
| PE/Cyanine7 anti-human TNF-α | MAb11 |
| Brilliant Violet 421 anti-human IL-2 | MQ1-17H12 |

* Bio Legend, Amsterdam, Netherlands

**Data analysis and statistics**

All data were collected using Microsoft Excel. Visualization and statistical analysis of all data was done in Studio (R version 4.2.2). Titers of binding RBD-, S and N-specific IgG as well as titers of neutralizing Abs were visualized in dot plots indicating average and ±1 standard deviation (SD). T-cell responses over time and compared to HNCs were visualized in boxplots unless mentioned otherwise in the legend. Box plots indicate median (middle line), 25th and 75th percentile (box), and the 5th and 95th percentile (whiskers). All data points, including outliers, are displayed, unless mentioned otherwise in the figure legend. Shapiro Wilk test was used to assess normality.

Statistics for serology and seroneutralization were performed on the log_10_ transformed average of two technical replicates, values below the detection range were recorded as 1/10^th^ the lower limit of quantitation (LLQ) and values above the detection range were recorded as upper limit of quantitation (ULQ). One-way analysis of variance (ANOVA) was utilized for longitudinal and cross-sectional comparisons of IgG titres, and neutralizing antibodies, across timepoints followed by pairwise two-tailed t-tests. Post hoc p-value correction was conducted using Bonferroni’s multiple-comparison correction method for all analyses.

For cellular immunity analyses, a quality control of minimum 500 live cells was used. CD4^+^ T-cell counts were performed by both an automated T-cell analysis at clinical site and during AIM analysis at laboratory site, T-cell counts showed a significant correlation (Pearson’s R = 0.56; P < 0.001). Non-parametric comparative Kruskal Wallis test were used for all independent activation induced marker (AIM) analyses on PBMCs and Wilcoxon signed rank test for paired and unpaired PBMC/AIM analyses. Post hoc p-value correction was conducted using Bonferroni’s multiple-comparison correction method for all analyses.

Pearson’s correlation on parametric data and Spearman correlation on nonparametric data were utilized to assess the relationship between serology and seroneutralization data, and all other T-cell markers. Distribution differences of the different cytokine combinations between the groups were performed using the nonparametric permutation test using SPICE, distributed by the National Institute of Allergy and Infectious diseases. One, two, or three stars in the figures indicate p < 0.05, p < 0.01, and p < 0.005, respectively.
